# Upper Limb Motor Improvement after TBI: Systematic Review of Interventions

**DOI:** 10.1101/2020.11.12.20214478

**Authors:** Sandeep K. Subramanian, Melinda A. Fountain, Ashley F. Hood, Monica Verduzco-Gutierrez

**Author notes:** Corresponding Author: Sandeep K Subramanian, Ph.D, B.P.Th, Assistant Professor, Department of Physical Therapy, School of Health Professions, UT Health San Antonio, San Antonio, Texas, USA 78229.

## Abstract

**Background:** Traumatic Brain Injury (TBI) is a leading cause of adult morbidity and mortality. Individuals with TBI have impairments in both cognitive and motor domains. Motor improvements post-TBI are attributable to adaptive neuroplasticity and motor learning. Majority of the studies focus on remediation of balance and mobility issues. There is limited understanding on the use of interventions for upper limb (UL) motor improvements in this population.

**Objective:** We examined the evidence regarding the effectiveness of different interventions to augment UL motor improvement after a TBI.

**Methods:** We systematically examined the evidence published in English from 1990-2020. The modified Downs and Black checklist helped assess study quality (total score:28). Studies were classified as excellent:24-28, good:19-23, fair:14-18 and poor:≤13 in quality. Effect sizes helped quantify intervention effectiveness.

**Results:** Twenty-three studies were retrieved. Study quality was excellent(n=1), good(n=5) or fair(n=17). Interventions used included strategies to decrease muscle tone (n=6), constraint induced movement therapy (n=4), virtual reality gaming (n=5), noninvasive stimulation (n=3), arm motor ability training (n=1), stem-cell transplant (n=1); task-oriented training (n=2) and feedback provision (n=1). Motor impairment outcomes included Fugl-Meyer Assessment, Modified Ashworth Scale, and kinematic outcomes (error and movement straightness). Activity limitation outcomes included Wolf Motor Function Test and Motor Activity Log. Effect sizes for majority of the interventions ranged from medium(0.5-0.79) to large(≥0.8). Only ten studies included retention testing.

**Conclusion:** There is preliminary evidence that using some interventions may enhance UL motor improvement after a TBI. Answers to emergent questions can help select the most appropriate interventions in this population.

## Introduction

Traumatic Brain Injury (TBI) is a major worldwide cause of morbidity and mortality.^1^ In the USA, recent reports indicate 2.87 million TBI related visits to the emergency room,^2^ with epidemiological data suggesting males being more affected than females.^3^ Common causes of TBI include falls, motor vehicle accidents and/or assaults.^4^ The available total annual cost estimates for TBI range from $56-$221 billion.^5^ Individuals sustaining a TBI may face cognitive,^6^ behavioral^7^ and communication difficulties^8^ lasting from few days post-injury to the rest of their lives.^9^ Additionally, a TBI causes sensorimotor impairments to the upper (UL) and lower limbs (LL).

Motor impairments include abnormal posture, altered muscle tone, paresis, reappearance of primitive and tonic reflexes, ataxia, decreased balance, and lack of coordinated movement.^10^ Individuals continue to have limited performance of activities of daily living, especially those relying on coordinated movements and UL muscle strength after a TBI. Persistent UL impairments and limitations in performance of daily life activities impact functional independence in this population.^11^

Motor improvements post-TBI are attributable in part to motor learning and adaptive neuroplasticity.^12^ Provision of rehabilitation benefits motor recovery by focusing on performing accurate repetitions of desired movement,^13^ is an integral part of motor learning and promotes adaptive neuroplasticity.^14^ Recent guidelines stress application of task-specific and intensive repetitive practice of functional reaching and activities including fine motor coordination.^15^

There is a need to identify the most effective and pertinent interventions with a focus on remediation of impairments and activity limitations in this population.^16^ To date, research has focused primarily on cognitive impairments and gait limitations, with less focus on UL issues.^17, 18^ This is an important topic, given that the UL issues are more diffuse and tend to be long standing in individuals post-TBI.^19^ Previous studies have identified deficits in UL functioning including impaired timing, reduced reach accuracy and grasping ability.^20^ Improving UL motor functioning helps boost the ability of individuals with TBI to perform activities of daily living such as dress, wash clothes, cook and groom.^21^ Enhanced UL functioning also enables better community reintegration post-TBI. For e.g., improving ability to drive helps commute to work and ability to be competitively employed, volunteer and/or attend school.^22, 23^

Our study objective was to systematically review the available literature focusing on rehabilitation of the UL, in individuals sustaining a TBI. Better identification of useful interventions can help select the best options to be used in the clinic and contribute to evidence-based practice. Our question in the Population, Intervention, Comparison and Outcome (PICO)^24^ format was, “In individuals sustaining a TBI, does provision of rehabilitation interventions augment UL motor improvement post-intervention compared to pre-intervention?” Preliminary results have previously appeared as an abstract.^25^

## Methods

### Systematic Literature Review

We performed a systematic search of the literature using Medline, Google Scholar, ISI Web of Science, Science Direct, and CINAHL. A Health Sciences Library Liaison helped formulate appropriate search strategies. Keywords and MeSH terms used included “traumatic brain injury”, “head injury”, “concussion”, “arm”, “upper limb”, “upper extremity”, “rehabilitation”, “intervention”, “motor recovery”, “impairment”, “activity limitation” and “motor improvement”. We used the terms “AND” and “OR” to combine keywords. Searches involved additional limits to restrict the articles to the English language literature published from January 1990 through August 2020, human species, and adult participants. Inclusion criteria were i) exposure to or provision of rehabilitation interventions and ii) assessment of motor impairment and/or limitations in activities of daily living using the UL. Exclusion criteria were i) studies focusing on effects of provision of only cognitive rehabilitation; ii) rehabilitation focusing exclusively on LL outcomes or iii) review articles, single case studies and expert opinion articles. We reviewed the reference lists of retrieved studies to identify additional relevant citations. We also checked the excluded reviews to identify any pertinent citations.

### Data Abstraction and Analysis

Retrieved articles were grouped according to the intervention used. We developed a data abstraction form to extract data from the selected articles. Data were initially extracted by MKF and AFH. The first author (SKS) then verified that all relevant data were obtained from the selected articles. The extracted data included details about chronicity, type of UL intervention, outcome of intervention and results of the study.

We quantified the effectiveness of interventions using estimates of effect sizes.^26^ When pre, post and retentions scores were available, effect sizes were calculated as the mean post-pre/SDpre values or mean retention - pre/ SDpre values. In case only change scores were reported, we used the ratio of the mean change score to the variability in change scores. Effect sizes (ES) ranging from 0.2-0.49, 0.5-0.79 and ≥0.8 were interpreted as small, medium, and large, respectively.^27^ We assessed the quality of the selected articles using the modified version^28^ of the Downs and Black checklist.^29^

The Downs and Black checklist is a reliable and valid assessment.^30^ It can be used to assess the quality of both randomized and non-randomized study designs. The total scores of this assessment and PEDro scale are highly correlated in studies involving individuals with brain injuries.^31, 32^ Scores on the Modified Downs and Black checklist were rated as “excellent” (score 24-28), “good” (score 19-23), “fair” (score 14-18), or “poor” (score ≤ 13).^33^ The quality of each study was independently evaluated by AFH and MKF, with discrepancies, if any, resolved by SKS.

## Results

A total of 140 citations were identified through database searches (Figure 1). After removing duplicates, 120 citations were screened, of which 90 were excluded. Of the 30 full text articles assessed for eligibility, we excluded seven studies, as they were reviews and/or expert opinions. Twenty-three articles were included in the qualitative synthesis. The different interventions used included those to reduce muscle tone (n=6), constraint induced therapy (n=4), virtual reality based gaming (n=5), non-invasive stimulation (n=3) [including neuromuscular electrical stimulation (n=1) and transcranial direct current stimulation (n=2)], Arm Motor Ability training (n=1), use of stem cells (n=1), goal oriented task-specific practice (n=1), feedback provision (n=1) and forced use therapy (n=1). The average (95% CI) age of participants was 36.4 (29.1 to 43.6) years. Brief highlights of the studies are presented below, with details in the accompanying tables. The scoring for the modified D&B checklist for each individual study is available in Supplementary table 1.

**Figure 1:**
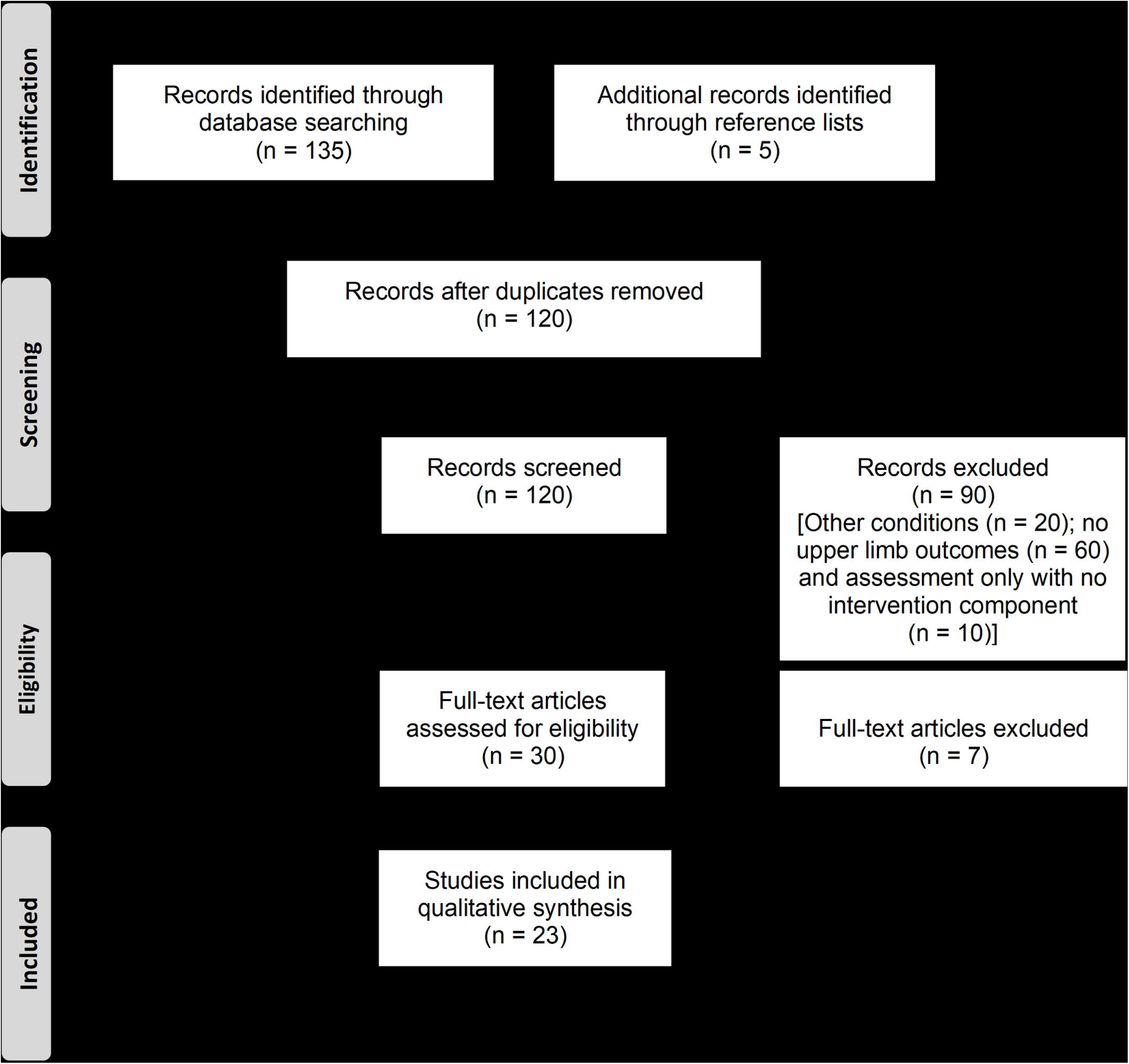
Preferred Reporting Items for Systematic Reviews and Meta-Analyses (PRISMA) flow

### A. Interventions to reduce muscle tone

We found six studies (quality ranging from fair to good) that examined the effects of different interventions on UL muscle tone (Table 1). Studies investigated the effects of different interventions on muscle tone reduction including provision of Botulinum toxin A injections, oral medication, serial casting soft splinting and acupuncture.

**Table 1.** Interventions to reduce muscle tone in the upper limbs

| *Study;*<br>*Sample*<br>*size (n)*<br>*and*<br>*Down's*<br>*and Black*<br>*score* | *Chronicity and*<br>*severity of injury* | *Intervention* | *Rehabilitation*<br>*provided/Dose* | *Outcomes* | *Timing of*<br>*assessment* | *Results* |
| --- | --- | --- | --- | --- | --- | --- |
| Yablon et al 1996;<br>n = 21<br>DBS 17<br>(fair) | 9 acute and 12 chronic.<br>Majority had severe injuries (GCS ≤8) | 20-40 units of Botulinum Toxin A injected under EMG guidance. | ROM therapy, casting and/or modalities provided as required. | <u>Wrist:</u><br>Modified Ashworth's scale for flexors and passive ROM using goniometry. | Baseline and 2-4 weeks after injections | <u>Acute TBI:</u><br>Decrease in tone measured by Modified Ashworth Scale; ES = -2.83<br>All participants had a lower MAS score (1-2 points), which is MCID<br>Improvement in wrist extension ROM; ES = 1.52.<br><u>Chronic TBI:</u><br>Decrease in tone measured by Modified Ashworth Scale; ES = -1.63<br>11 out of 12 participants had a lower MAS score (1-2 points), which is MCID<br>Improvement in wrist extension ROM; ES = 1.74 |
| Pavesi et al. 1998;<br>n = 6<br>DBS 14<br>(fair) | Spasticity present for 4-6 months post-injury | 20-40 units of Botox; injected under EMG guidance. | After injections, casting was provided. | <u>Wrist:</u><br>Modified Ashworth's scale for flexors and passive ROM using goniometry. | Baseline and 4 weeks after injection | Decrease in tone measured by Modified Ashworth Scale; ES = -2.38<br>All participants had a lower MAS score (1-2 points), which is MCID |
|  | Severe injuries (GCS $\leq 8$) |  |  |  |  | Improvement in wrist extension ROM; ES = 2.11 |
| Meythaler et al 2001;<br>n = 17 (8 TBI)<br>DBS 22 (good) | Spasticity present for at least 6 mos. before study participation<br>Initial injury severity level information missing | Oral Tizanidine for a maximum of 36 mg/d or dose tolerated for 8 weeks followed by placebo or vice versa. | No information provided | <u>Muscle tone:</u><br>Combined Original Ashworth's scale score for shoulder abductors, elbow muscles and wrist extensors.<br><u>Activity limitations:</u><br>Functional Independence Measure (FIM) and Craig Hospital Assessment and Reporting Technique. | Baseline and 4 weeks after start of medication.<br>Retention assessment only when active drug administered | Greater reduction in tone with Tizanidine (ES = -0.36) compared to placebo (ES = -0.23; $p < 0.05$).<br>Effect not retained.<br>No improvements noted in activity limitations. |
| Moseley et al 2006;<br>n = 26<br>DBS 22 (good) | Duration since injury $\leq 6$ months.<br>Severe injuries (GCS $\leq 5$) | Serial casting (n=14) or positioning (n=12) | *Serial casting group*<br>Elbows stretched in an extended position for 14 days.<br>Progressive increase of stretch range after first 7 days.<br>*Positioning group*<br>Passive stretch applied for one hour/day; 5-7 days/week. | <u>Primary:</u><br>Torque controlled elbow extension.<br><u>Secondary:</u><br>Tone assessed using Modified Tardieu Scale;<br>Function assessed using the Test Évaluant la Performance des Membres Supérieurs des Personnes Âgées (TEMPA). | Baseline, immediately after cast removal, and at one month retention.<br>An additional assessment conducted one day after cast removal for the primary outcome. | *Primary Outcome*<br>Improved elbow extension range by 22° after serial casting for 2 weeks; ES = 1.31<br>One day after cast removal, gain in elbow range decreased to 15 degrees; ES = 1.17<br>Gain of 11° elbow extension range maintained 1 day after removal of stretch; ES = 0.29<br>Change was not maintained at retention |
|  |  |  | • Stretch position maintained using sandbags, slings or splints.<br>• Both groups also received additional therapy designed to improve individual motor skill (15 minutes/day, five days/week). |  |  | *Secondary Outcomes*<br>• Tone reduced immediately after serial casting compared to positioning; ES = -1.19.<br>• Reduction in tone not maintained at retention.<br>• Both groups improved on TEMPA; no between group differences seen. |
| Thibaut et al 2015;<br>N = 17 (7 TBI)<br>DBS 20 (good) | • Duration since injury $\geq 3$ months.<br>• Severe injuries – 6 participants minimally conscious and 11 in a vegetative/unresponsive wakeful state. | Participants received three different one-hour long interventions in a randomized crossover fashion. | • The interventions consisted of<br>a) stretching (30 mins) and splint (30 mins);<br>b) splint (30 mins) and no treatment (30 mins);<br>c) manual stretching (30 mins) and no treatment (30 mins). | Modified Ashworth Scale for finger flexors and passively measured distance from thumb to fingers. | Baseline, immediately after treatment and 60 minutes after treatment (retention). | *Stretching and splinting:*<br><u>Tone:</u><br>• Immediate decrease in Modified Ashworth Scale score after intervention; ES = -1.08<br>• Overall change in MAS score by 1 point, which is MCID.<br>• Reduction maintained at retention; ES = -0.54<br><u>Hand opening distance:</u><br>• Increased hand opening at after intervention; ES = 0.55<br>• Change not maintained at retention.<br>*Splinting only*<br><u>Tone:</u><br>• Immediate decrease in Modified Ashworth Scale score after intervention; ES = -0.68 |
|  |  |  |  |  |  | • Overall change in MAS score by 1 point, which is MCID.<br>• Reduction maintained at retention; ES = -0.33<br><u>Hand opening distance:</u><br>• Increased hand opening at POST compared to PRE; ES = 0.75<br>• Change not maintained at RET<br>*Stretching only*<br>• No significant changes seen at POST or RET. |
| Matsumoto - Miyazaki et al. 2016; N = 11 DBS 20 (good) | • Duration since injury $\geq 8$ months.<br>• Severe injuries – 7 participants minimally conscious and 4 in a vegetative/unresponsive wakeful state. | Acupuncture or sham stimulation provided. one week apart in a crossover randomized manner. | Stimulation provided on the face, dorsum of the hand near the second metacarpal and anterior aspect of leg near the tibialis anterior muscle.<br>Stimulation provided. for a total of 10 minutes. | Abductor Pollicis Brevis F/M ratio.<br>16 F waves recorded for the Abductor Pollicis Brevis muscle with stimulation provided at the median nerve. | Baseline, immediately after removal of the needle (10 minutes) and at 20 minutes | *Acupuncture*<br>• Decrease in F/M ratio after immediately after intervention; ES = -0.73<br>• Change maintained at retention; ES = -0.70.<br>*Sham stimulation*<br>• No change in F/M ratio seen post- stimulation or at retention. |

DBS: Downs and Black Checklist Score; TBI: Traumatic Brain Injury; GCS: Glasgow Coma Scale; EMG: Electromyography; ROM: Range of Motion; ABI: Acquired Brain Injury; ES: Effect Size; MCID : Minimal Clinically Important Difference

Two studies ^34, 35^ investigated the effects of Botulinum toxin A injections on wrist flexor muscle tone in 27 individuals post-TBI (18 males, 9 females) with moderate-to-severe muscle tone. Botulinum toxin A injections were delivered to target muscles under EMG guidance. Changes in muscle tone (quantified using Modified Ashworth’s Scale) and wrist extension range of motion (measured using goniometry) helped assess the effects of the injections. Muscle tone decreased and wrist extension range increased following Botulinum toxin A injections (ES>0.8).

Meythaler et al^36^ assessed the effects of oral tizanidine administration on UL muscle tone in 17 individuals (14 males, 3 females) with acquired brain injuries (ABIs; TBI: n=8, stroke: n=9). They administered either tizanidine or placebo in a crossover fashion for 6 weeks, tapered the drug for one week and then switched over to other medication after one more week. Oral tizanidine decreased muscle tone (assessed using Original Ashworth’s scale) on the affected side immediately after treatment (ES= −0.36) with no retention at 6 weeks (ES= −0.1).

Moseley et al^37^ recruited 26 individuals (23 males, 3 females) post-TBI with elbow flexion contracture, and randomized them into two groups (n=13/group). One group received serial casting, and the other received static positioning. Serial casting increased elbow range by 22^°^ over static positioning immediately post-intervention (ES=1.85). One day after cast removal, elbow range gain decreased to 15^°^ in the serial casting group (ES=1.17), which further decreased to 11^°^ after 2 weeks post-intervention.

Thibaut et al^38^ randomized 17 participants (10 males, 7 females) with ABIs (TBI: n=7, stroke: n=10) to receive either soft splinting, 30 minutes of manual stretching, or no treatment. Provision of soft splinting resulted in increased hand opening ability (2.39 cm of major-palm distance, ES=0.55). Additionally, soft splinting and manual stretching decreased finger flexor muscle tone after 30 minutes of treatment (ES= −0.53 and −0.55).

Matsumoto et al^39^ assessed effects of acupuncture provision on UL muscle tone in 11 unconscious or minimally conscious males. They used a crossover study design providing acupuncture or no treatment, separated by one week. Acupuncture provision reduced the F/M ratio at the end of treatment (ES= −0.73) and was retained 10 minutes later (ES= −0.7).

### B. Constraint Induced Movement Therapy (CIMT)

We found four studies (fair quality; Table 2) that assessed the effects of CIMT on UL impairment, activity, and self-reported UL use levels. Page and Levine,^40^ in a case series involving three participants (2 males, 1 female) post-TBI, constrained the less-affected side (5hours/day; 10 weeks). All participants improved UL activity performance measured using the Wolf Motor Function Test - Functional Ability Scale (WMFT; ES=3.0) and Action Research Arm Test (ES=1.78). Additionally, participants also improved the amount and quality of self-perceived use, assessed using the Motor Activity Log (MAL).

**Table 2.** Use of Constraint Induced Movement Therapy (CIMT)

| *Study;*<br>*Sample*<br>*size (n)*<br>*and*<br>*Down's*<br>*and Black*<br>*score* | *Chronicity and*<br>*severity of injury* | *Intervention* | *Rehabilitation*<br>*provided/Dose* | *Outcomes* | *Timing of*<br>*assessment* | *Results* |
| --- | --- | --- | --- | --- | --- | --- |
| Page and Levine 2003;<br>n = 3<br>DBS 15<br>(fair) | • All participants had chronic injuries<br>• Initial injury severity level information missing | Modified CIMT.<br>Mitt worn on the less-affected side | All individuals wore the mitt 5 days/week, for 5 hours/day for 10 weeks.<br>They also received three 30 mins long sessions/week of both PT and OT for 10 wks. | • Motor Activity Log Quality of Movement (MAL-QoM) and Amount of Use (MAL-AoU) scales,<br>• Wolf Motor Function Test – Functional Assessment Scale (WMFT-FAS) and<br>• Action Research Arm Test (ARAT) | • Baseline and after intervention completion. | • Improvements in MAL QoM and AoU scales above MCID levels.<br>• Improved WMFT – FAS (ES = 3.0) and ARAT scores (ES = 1.78) after intervention completion. |
| Shaw et al 2005;<br>n = 22<br>DBS 18<br>(fair) | • All participants had chronic injuries<br>• Initial injury severity level information missing | Traditional CIMT<br>The mitt was worn 90% of waking hours on the less-affected side. | All individuals practiced UL activities for 6 hrs/day, 5 days/wk for 2 wks.<br>Participants were encouraged to perform better and explicit verbal feedback on small improvements was provided. | FMA, WMFT-FAS, and MAL-QoM scale. | • All assessments completed at baseline and after intervention completion. Only MAL-QoM additionally assessed at retention (one mo. and 2 yrs). | <u>FMA scores</u><br>• Improved (ES = 1.4) scores after intervention completion.<br>• Greater change seen in those with mild-to-moderate (ES = 1.4) and severe (ES = 1.6) impairment compared to moderate impairment (ES = 1.0). |
|  |  |  |  |  |  | <u>WMFT scores</u><br>• Improved WMFT-FAS (ES = 0.7) after intervention completion.<br>• Greater change seen in those with moderate impairment (ES = 1.3) compared to mild-to-moderate (ES = 0.6) and severe (ES = 0.5) impairment<br><u>MAL scores</u><br>• MAL-QoM scores improved at the end of the intervention (ES = 2.1) with change retained at 1 month (ES = 2.1) and 2 years (ES = 1.3). These changes exceeded published MCID levels.<br>• Compared to those with severe impairment, greater change seen immediately post treatment in those with and at 1-mo retention (mild-to-moderate impairment; ES = 2.4) and moderate impairment ;ES = 3.7). |
| Morris et al. 2006; n= 29<br>DBS 17 (fair) | • All participants had chronic injuries<br>• Initial injury severity level information missing | Traditional CIMT<br>The mitt was worn 90% of waking hours on the less-affected side. | All individuals were involved in performance of UL activities for 6 hours/day, 5 days/week for 2 weeks | • FMA, WMFT-FAS, WMFT time to complete activities and MAL QoM and AoU scales.<br>• Visual attention, task-switching and global | • Baseline and after intervention completion.<br>• Cognitive factors were assessed as part of inclusion criteria | <u>FMA scores</u><br>• The intervention led to improvements in FMA scores (ES = 1.5)<br><u>WMFT scores</u><br>• The intervention led to improvements in WMFT FAS scores (ES = 0.4) and |
|  |  |  |  | cognition were also assessed |  | time to complete activities (ES = 0.2).<br>• Individuals were 22.6% times faster in completing activities, which is at MCID levels.<br>*<u>MAL scores</u>*<br>• The intervention led to improvements in and MAL QoM (ES = 2.1) and AoU (ES = 1.7) scores.<br>• These changes exceeded published MCID levels.<br>• Increase in self -perceived arm use correlated with better global cognition, visual attention and task-switching. |
| Cho et al. 2005;<br>n = 9 (3 TBI)<br>DBS 14 (fair) | • Injury sustained $\geq 12$ weeks before study participation.<br>• Initial injury severity level information missing | Traditional CIMT with splints that prevented contact of thumb and index finger on the less-affected side<br>The three participants with TBI for two, three or five weeks. | The individuals continued with whatever therapy was previously prescribed.<br>Exact details of dose in terms of time spent or numbers of repetitions missing. | • Perdue Pegboard Test | • Weekly, scores on the Perdue Pegboard test recorded and scoring stopped when no change was seen for 3 consecutive weeks | • Wearing splint resulted in improved performance on the test (ES = 1.31). |

DBS: Downs and Black Checklist Score; TBI: Traumatic Brain Injury; ABI: Acquired Brain Injury; FMA: Fugl Meyer Assessment; WMFT-FAS: Wolf Motor Function Test – Functional Assessment Scale; MAL-QoM : Motor Activity Log Quality of Movement; MAL-AoU: Motor Activity Log Amount of Use; ES: Effect Size; MCID: Minimal Clinically Important Difference

Two studies examined the effects of CIMT on participants with chronic TBI. In both studies, participants wore the mitt on the less-affected limb for 90% of waking hours. All participants (n=22; 14 males, 8 females) in the first study by Shaw et al^41^ decreased UL motor impairment measured using the Fugl-Meyer Assessment (FMA; ES=1.4) and improved in performance of daily life activities (measured using WMFT; ES=0.7) immediately after treatment. Participants also reported an increase in self-perceived quality of movement immediately after the intervention (ES=2.1), which was retained at one month (ES=2.1) and at two years post-intervention (ES=1.3).

Participants in the other study by Morris et al.^42^ (n=29; 19 males, 10 females) similarly had better scores on the FMA (ES=1.5) and WMFT (ES=0.4). Participants reported an increase in the amount (ES=1.7) and quality (ES=2.1) of self-perceived UL use after the intervention. Participants reporting better use of the more-affected UL had better global cognition (assessed using the Mini-Mental Scale) and visual attention and task switching (measuring using the Trail Making Tests A and B).

Cho et al^43^ examined the effects of CIMT on fine motor function of the hand in 9 participants (8 males, 1 female) with ABIs (TBI: n=3, stroke: n=6). The less-affected side was partially constrained with an opposition restriction splint that blocked use of the thumb and index finger. All participants were evaluated weekly using the Perdue Pegboard test, until no change was seen in three consecutive assessments. Constraining the less-affected side resulted in improved performance on the pegboard test (ES=1.31).

### C. Virtual Reality Gaming

We found five studies (fair to good quality; Table 3) that assessed the effects of VR on motor performance outcomes and UL functional outcomes.

**Table 3.**
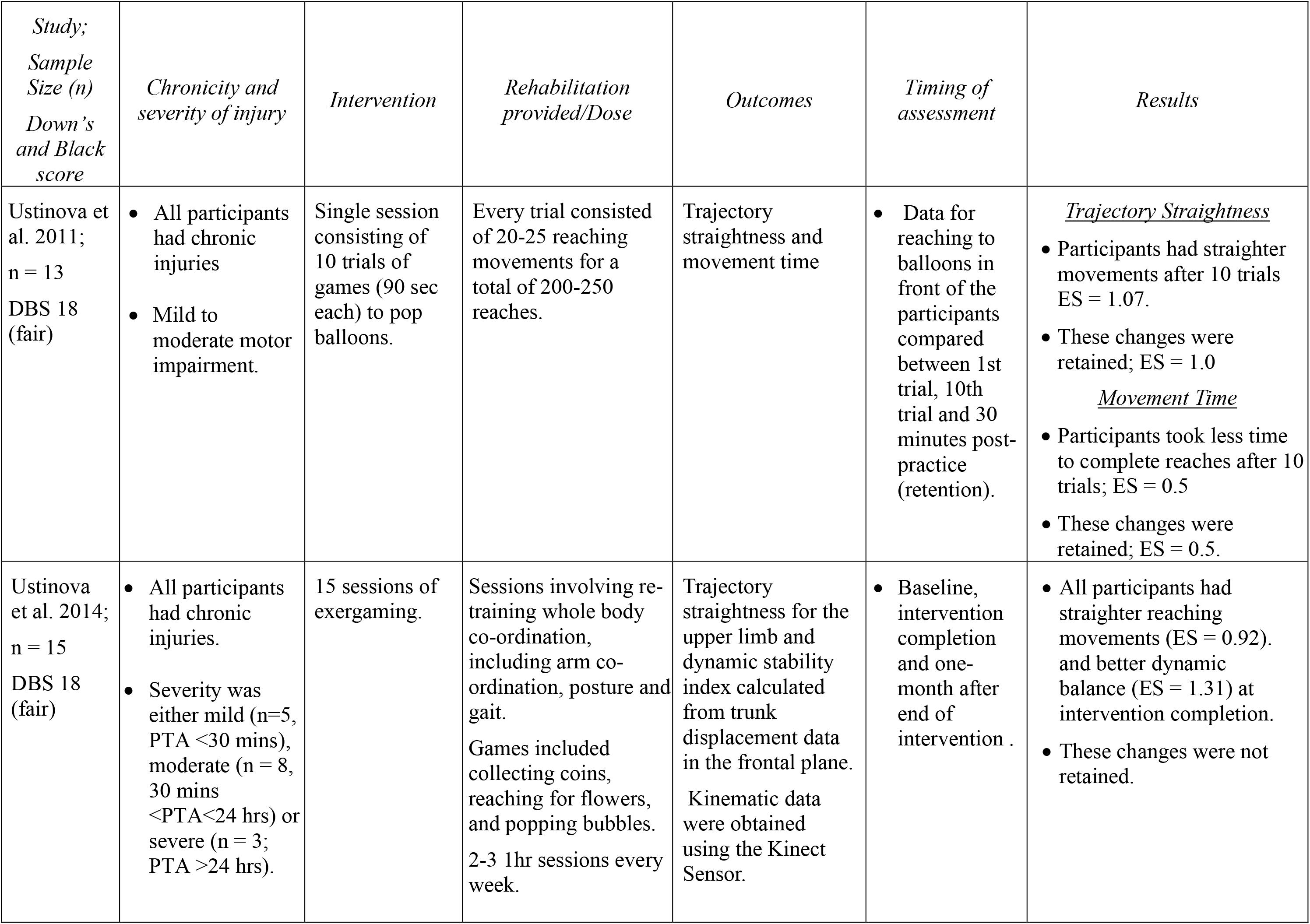
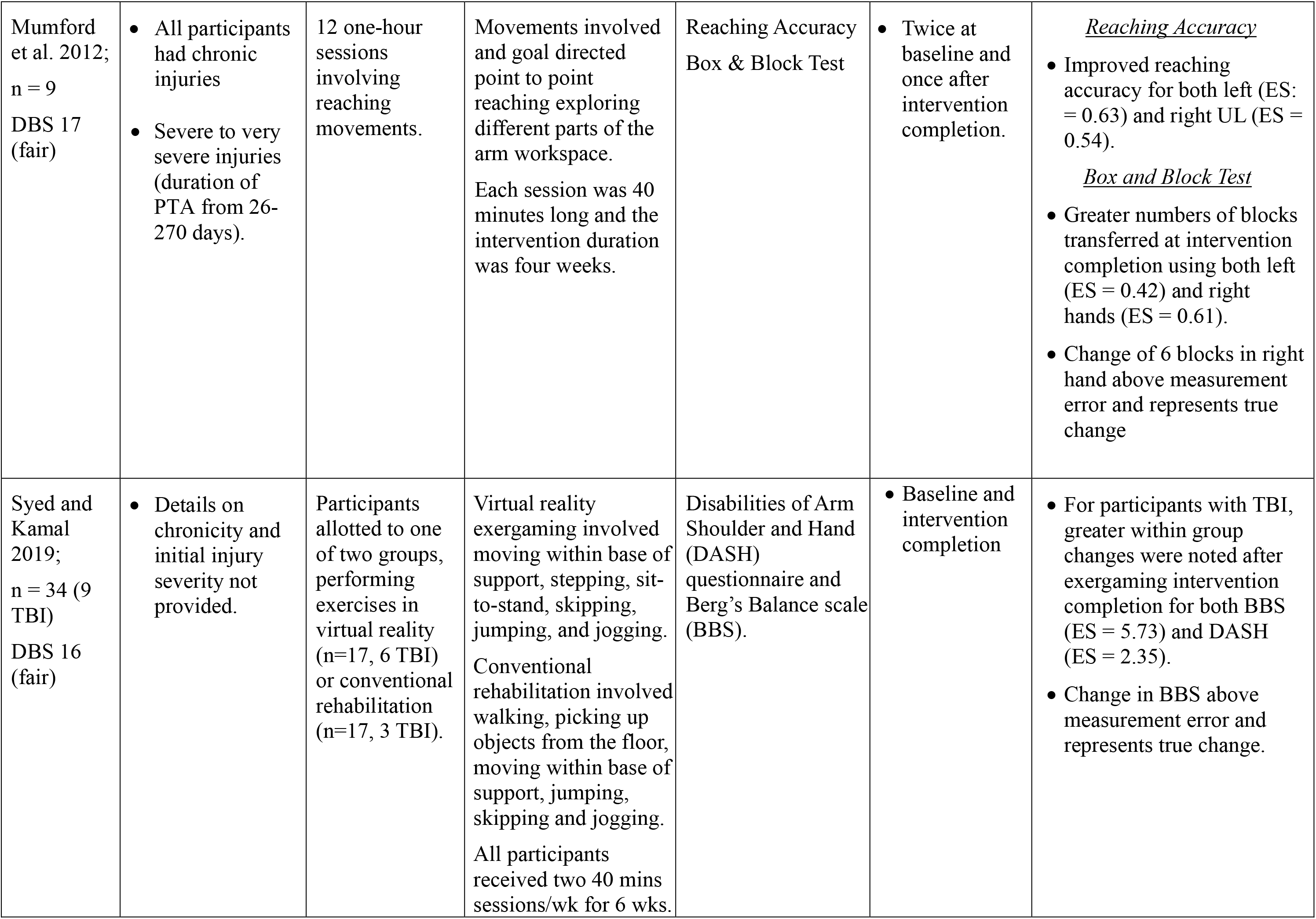
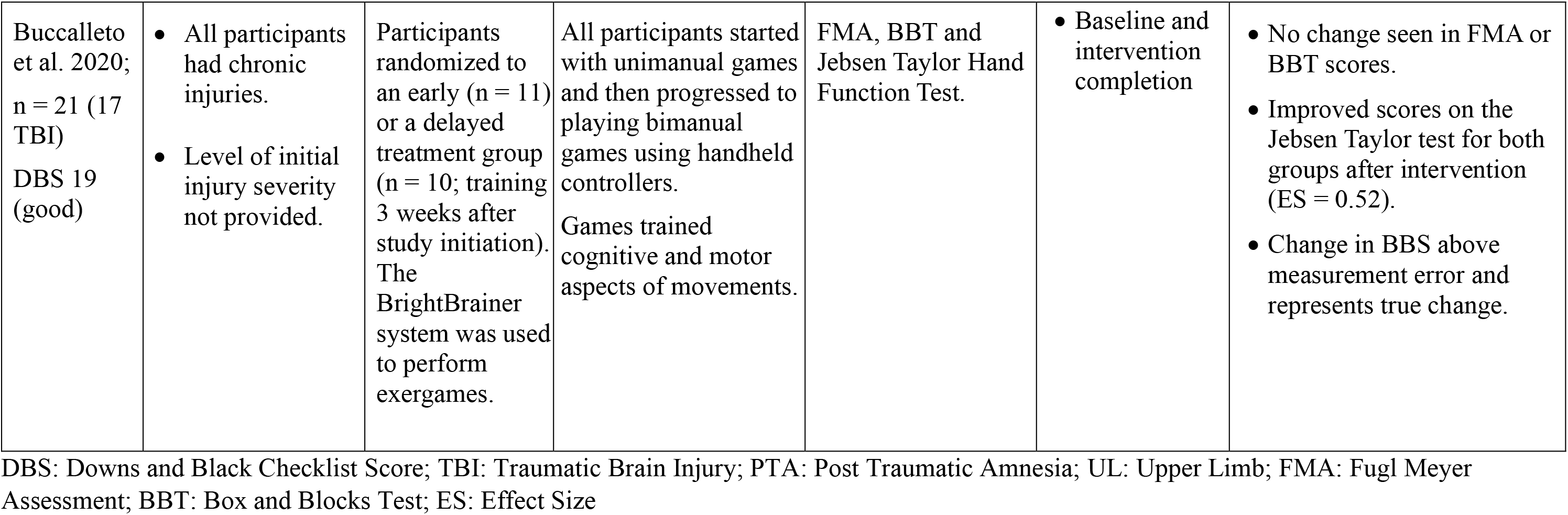
Use of Virtual Reality gaming

In two studies, Ustinova and colleagues examined the effects of task-practice of reaching movements from a standing position. In the first study,^44^ 13 participants post-TBI (6 males, 7 females) practiced 10 trials of reaching movements. Movements were recorded using a motion analysis system. At the end of 10 trials, participants reached faster to the targets (ES=0.54), and further improved (ES=0.74) at retention testing (30 minutes post-intervention). The participants also had straighter reaching movements (ES= −1.07), which were retained (ES= −0.97).

The second study,^45^ examined the effects of multiple sessions of playing games on balance, reaching and co-ordination. Participants (n=15; 10 males, 5 females) with chronic TBI played games for 15 sessions (thrice weekly). All participants were assessed at baseline, after practice and one-month post-practice. Dynamic balance (ES=1.33) and reaching movement straightness (ES= −1.16) improved after practice and at one month, these changes were retained.

Mumford et al^46^ examined the effects of repetitive practice of unimanual and bimanual UL movements in nine individuals (5 males, 4 females) with severe chronic TBI. Assessments included kinematic measures of reaching as well as the number of blocks transferred on the Box and Blocks Test (BBT). After training, all participants had more accurate movements (ES=0.62) and transferred more blocks (ES=0.42).

Syed and Kamal^47^ assessed the effects of VR-based gaming on 34 individuals with a variety of neurological disorders (26 males, 8 females) including TBIs (n=9). Participants received 12 sessions of either VR-based training (n=17) or conventional training (n=17). Both groups improved after training with greater changes noted with VR-based (p<0.001) compared to conventional training (p<0.05) for both balance (assessed using Berg Balance Scale, BBS) and self-reported UL ADL performance [assessed using Disabilities of the Arm, Shoulder and Hand (DASH) questionnaire). When results were compared only for the participants post-TBI, greater within group changes were noted after VR-based training compared to before for BBS (ES=5.73) and DASH (ES=2.35).

In another study, Buccellato et al^48^ examined the effects of VR-based gaming on a group of 21 participants (15 males, 6 females) with ABIs (TBI: n=13, stroke: n=4, a combination of stroke + TBI: n=4). Participants were randomized to an early treatment group (n=11) or a delayed treatment group (began training 3 weeks after study initiation; n=10). The effects of this system on UL function, dexterity and activity performance was assessed using the FMA, BBT and Jebsen Taylor Hand Function test, respectively. Early or delayed training did not result in improved function or dexterity. However, activity performance was improved (ES=0.52).

### D. Non-invasive Stimulation

We found 3 studies (fair quality) that assessed the effects of non-invasive stimulation including use of neuromuscular electrical stimulation (NMES; one study; Table 4A) and transcranial direct current stimulation (tDCS; two studies; Table 4B) on UL motor improvement after TBI.

**Table 4.** Use of Noninvasive Stimulation and Arm Ability Training

| *Study;*<br>*Sample size*<br>*(n),*<br>*Down's and*<br>*Black score* | *Chronicity and*<br>*severity of*<br>*injury* | *Intervention*<br>*details* | *Rehabilitation*<br>*provided/Dose* | *Outcomes* | *Timing of*<br>*assessment* | *Results* |
| --- | --- | --- | --- | --- | --- | --- |
| *A. Use of NeuroMuscular Electrical Stimulation* |  |  |  |  |  |  |
| Alon et al.<br>1998;<br>n = 20 (7<br>TBI)<br>DBS = 17<br>(fair) | All participants had chronic injuries<br>Initial injury severity level information missing | Provision of NMES enabling reciprocal finger flexion and extension along with grasp and release. | Forearm-hand splint with surface electrodes positioned over the wrist and hand muscles.<br>Pulses delivered in an interrupted mode; contraction and relaxation intervals: 3-7 secs<br>Daily average of 3.5 hrs stimulation for a total of 4 mos. | Resting postures, active, and passive ROM at the wrist and elbow joints assessed using goniometry at the beginning and end of the intervention | Baseline and after intervention completion | *Wrist Joint*<br>More extended resting posture (ES = 3.71) at the intervention completion.<br>Increased range of passive (ES = 2.69) as well as active extension (ES = 2.73)<br>*Elbow Joint*<br>More extended resting posture (ES = 4.09) at the intervention completion.<br>Increase in active elbow extension ROM (ES = 6.91) |
| *B. Use of Transcranial Direct Current Stimulation* |  |  |  |  |  |  |
| Kang et al<br>2012;<br>n = 9<br>DBS = 18<br>(fair) | Participants had either sub-acute (n = 4) or chronic (n = 5) injuries.<br>Initial injury severity | Real (2mA anodal) or sham stimulation | tDCS stimulation over DLPFC applied for 20 minutes.<br>Sham stimulation consisted of 1 min ramp up and ramp down. | Reaction time on Contrast reaction time task | Baseline, immediately after intervention completion, 3 hrs and 24 hrs after intervention completion. | Tendency (p = 0.056) to reduce reaction time after real compared to sham stimulation; ES = 0.89.<br>This change was not maintained 3 hours post stimulation (ES = 0.1). |
|  | level information missing. |  |  |  | MMSE scores assessed only at baseline. | However, better retention of change in reaction times 24 hours post stimulation (ES = 0.96) and this change correlated moderately with initial MMSE scores ($r = 0.67$) |
| Middleton et al 2014;<br>n= 5 (2 TBI)<br>DBS = 14 (fair) | Both participants had chronic injuries<br>Initial injury severity level information missing | Bihemispheric stimulation of 1.5 mA for 20 minutes.<br>Total of 24 sessions, sessions held thrice weekly. | Stimulation followed by intensive task-specific practice of UL gross and fine motor activities.<br>*Gross motor activities* - reaching for items on shelves, hitting a balloon with a racquet and simulating household chores.<br>*Fine motor activities* - flipping playing cards and manipulating small change. | Clinical: FMA and BBT<br>Kinematic: Movement straightness and speed assessed using the robotic manipulandum.<br>Assessments completed immediately after practice and at 6-month retention assessment. | Baseline, immediately after intervention completion, and 6 mos. after intervention completion | *In participants with TBI-*<br>Intervention led to better FMA scores (ES = 0.47), which was retained (ES = 0.42).<br>Change in FMA scores exceeded MCID levels.<br>All participants moved faster (ES = 0.37). At retention, participants continued to move faster (ES = 0.70). |
| C. Use of Arm Ability Training |  |  |  |  |  |  |
| Platz et al. 2001;<br>n = 60 (15 TBI)<br>DBS = 25 (excellent) | Participants had acute to early chronic injuries (3 wks – 24 mos.) post injury.<br>Initial injury severity level information missing | Participants randomized into groups to received Arm Ability Training, Ability Training with knowledge of results feedback or no Arm Ability Training (n=20 each). | Arm Ability Training included activities involving dexterity manipulation, aiming for targets and gripping objects of different sizes.<br>The knowledge of results feedback group got average feedback. | Clinical: Hand function evaluated using the Test Evaluant les Membres superieurs des Personnes Agees (TEMPA).<br>Kinematic: Movement time for aiming movements | Baseline, intervention completion and one year after intervention completion. | *TEMPA*<br>Participants who received Arm Ability Training took less time to complete the TEMPA (ES = 0.95) at intervention completion.<br>Changes were retained at one year (ES = 0.82).<br>*Kinematic outcomes* |
|  |  |  | Details on what exact intervention the no Arm Ability Training group received are missing. | performed using a stylus. |  | • Participants in the Arm Ability Training group had faster movements at intervention completion (ES = 0.73).<br>• No information provided about retention testing.<br>• Provision of feedback did not result in additional gains. |

DBS: Downs and Black Checklist Score; TBI: Traumatic Brain Injury; ABI: Acquired Brain Injury; ROM: Range of motion; MMSE: Mini Mental Scale Examination; FMA: Fugl Meyer Assessment; BBT: Box and Blocks Test; ES: Effect Size; MCID: Minimal Clinically Important Difference

Alon et al^49^ assessed the effects of provision of NMES enabling reciprocal finger flexion and extension along with grasp and release in 20 individuals (14 males, 6 females) with chronic ABIs (TBI: n=7, stroke: n=13). All participants received an average of 3.5 hrs. stimulation daily over the course of the intervention, which lasted for almost 4 months. All participants had a more extended posture at the elbow (ES=4.09) and wrist (ES=3.71) at rest at the end of the intervention. At the wrist, participants improved their range of passive extension (ES=2.69) as well as active flexion and extension (ES=2.73). At the elbow, active ROM of elbow movement increased (ES=6.91).

Kang and colleagues^50^ assessed the effects of 2mA anodal tDCS to the left dorsolateral prefrontal cortex on reaction time to an attention task. Nine individuals (8 males, 1 female) with chronic TBI participated and were randomized to receive active tDCS for 20 mins or sham stimulation after one week in a crossover fashion. Reaction time on a computerized timed task decreased after application of real tDCS vs sham stimulation at the end of the intervention (ES= - 0.89). However, this change was not maintained at the two retention assessments (3 hours and 24 hours after the end of stimulation).

Middleton et al^51^ examined the effects of bi-hemispheric stimulation followed by robotic training on five participants with ABIs (TBI; n=1, stroke: n=3, stroke + TBI: n=1). All participants performed strengthening and functional activities for a total of 40 minutes. Each participant received 1.5mA intensity concurrent stimulation for the first 15 minutes. Results for participants post-TBI (2 males) revealed improvements only in FMA scores (ES=0.47), which were retained (ES=0.42). Participants post-TBI also reached the targets faster at the end of the intervention (ES=0.37; assessed by the KINARM^©^ robotic device) and continued to improve 6 months later (ES=0.7).

### E. Arm Ability Training (AAT)

We found 1 study (excellent quality; Table 4C) that assessed the effects of AAT on motor performance outcomes and hand function. In this study by Platz and colleagues,^52^ 60 participants (36 males, 24 females) with ABI (TBI: n=15, stroke: n=45) were randomized into three groups: a control group, a group receiving AAT and a group receiving AAT + knowledge of results (KR) feedback (n=20 each). Activity performance was assessed using the time to complete the TEMPA (Test Evaluant les Membres Superieurs des Personnes Agees). Kinematic assessment of an aiming movement on a stylus between two targets was also conducted. Provision of AAT resulted in faster performance on the TEMPA (ES=0.95), which was retained one year later (ES=0.75). Participants receiving AAT also had faster aiming movements (ES=0.67). Providing KR feedback did not enhance task performance.

### F. Stem Cell Transplantation

We found 1 study (fair quality; Table 5A) assessing the effects of stem cell transplantation on motor impairment. This study^53^ examined the effects of provision of injection of mesenchymal stem cells derived from the umbilical cord. Forty participants (32 males, 8 females) with moderate to severe TBI were randomized to receive the injections or to a control group (n=20/group). The cells were injected into the sub-arachnoid space after lumbar puncture performed between the 3^rd^ and 4^th^ or 4^th^ and 5^th^ vertebrae. Motor impairment was assessed using the FMA at baseline and 6 months after the injection. The Funtional Independence Measure (FIM) helped quantify assistance in activity performance. The intervention group had significantly better improvement in FMA scores than the control group for both the UL (ES=1.38) and LL (ES=0.88) as well as FIM scores (ES=1.17).

**Table 5:** Use of Stem cells and other interventions

| *Study;*<br>*Sample size*<br>*(n),*<br>*Down's and*<br>*Black score* | *Chronicity and*<br>*severity of*<br>*injury* | *Intervention*<br>*details* | *Rehabilitation*<br>*provided/Dose* | *Outcomes* | *Timing of*<br>*assessment* | *Results* |
| --- | --- | --- | --- | --- | --- | --- |
| *A. Stem cell transplantation* |  |  |  |  |  |  |
| Wang et al. 2013;<br>n = 40<br>DBS = 18<br>(fair) | All participants had chronic injuries<br>All participants had severe injuries (mean GCS score of 7). | Participants were randomized into two groups to receive stem cells or a control group.<br>The stem cell group received an injection of umbilical cord mesenchymal stem cells.<br>2ml of stem cell suspension (containing $1 \times 10^7$ stem cells) injected into subarachnoid space between lumbar vertebrae 3 and 4 or 4 and 5. | Details on whether the intervention or control group received any form of rehabilitation are missing. | FMA and FIM. | Baseline and 6 mos. post injection | Significant improvement in upper (ES= 1.38) and lower limbs (ES = 0 .88) FMA scores as well as FIM scores (ES = 1.17) for the intervention group.<br>No change seen in control group. |
| *B. Other Interventions* |  |  |  |  |  |  |
| Sietsema et al. 1993;<br>n = 20<br>DBS = 18<br>(fair) | All participants had chronic injuries | Computer game involving reaching from a seated position or rote arm reaching. | Initially, NDT based intervention was provided.<br>Participants performed rote reaching exercises | Reaching distance between hip and wrist measured using a motion capture system | Baseline and after 10 trials in each condition | Greater reaching distance over the 10 trials of playing the game compared to rote reaching (ES = 1.41). |
|  | All participants had mild to moderate injuries (RLA staging score IV or V). |  | or played a computer game.<br>The game involved repeating a sequence of flashing lights and sounds by pressing certain buttons.<br>The order of playing the game or rote arm reaching was counterbalanced. |  |  |  |
| Croce et al. 1996;<br>n = 51<br>DBS = 18 (fair) | All participants had chronic injuries<br>All participants had moderate injuries (GCS score: 8 - 12). | Practice of an anticipation task that involved pressing a button when they saw the last light. | Participants divided into 4 groups depending upon when they received Knowledge of Results (KR) feedback into a no KR group (n = 12), 100% KR (n = 14), summary KR (after every 5 trials; n = 13) and average KR (average information for all 5 trials; n = 12)<br>Individuals performed 60 acquisition trials (12 blocks of 5 trials each). | Absolute error in terms of timing of responses. | Assessments conducted at intervention completion, ten minutes after the last acquisition trial, (immediate retention) and 24 hrs following the last acquisition trial (late retention). | Provision of KR better than no KR.<br>*100% KR*<br>Effective immediately after practice; ES = 0.96.<br>Effects reduced at immediate retention testing; ES = 0.67 and at late retention; ES = 0.37<br>*Summary KR*<br>Beneficial immediately after practice; ES = 0.97.<br>Changes maintained at immediate and late retention periods; ES = 1.21.<br>*Average KR*<br>Beneficial immediately after practice; ES = 0.95.<br>Changes maintained at early retention; ES = 1.02; but declined at late retention; ES = 0.77. |
| Sterr and Freigvogel, 2003;<br>n = 13 (11 TBI)<br>DBS = 18 (fair) | • All participants had chronic injuries<br>• Initial injury severity level information missing | Forced use therapy. | All participants initially received OT for 90 minutes for 4 weeks in phase A.<br>This was followed by forced use therapy involving principles of shaping for another 4 weeks in phase B.<br>The participants practiced 4-10 tasks in each session. | The Frenchay Arm Test; MAL AoU, MAL QoM as well as WMFT-FAS | • At the end of phases A and B, and one month after the end of Phase B. | *Immediately at the end of Phase B:*<br>• Significant improvements seen in Frenchay Arm Test scores (ES = 0.72), MAL AoU (ES = 2.38), MAL QoM (ES = 1.98) and WMFT-FAS (ES = 1.76).<br>• Change in MAL AoU and QoM scores at MCID level.<br>*Retention at 4 weeks post practice:*<br>• Changes were not retained compared to end of Phase A in any of the 4 clinical outcomes. |

DBS: Downs and Black Checklist Score; GCS: Glasgow Coma Scale; FMA: Fugl Meyer Assessment; FIM: Functional Independence Measure; ABI: Acquired Brain Injury; KR: Knowledge of Results; WMFT-FAS: Wolf Motor Function Test – Functional Assessment Scale; OT: Occupational Therapy; MAL-QoM : Motor Activity Log Quality of Movement; MAL-AoU: Motor Activity Log Amount of Use; ES: Effect Size ES: Effect Size; MCID: Minimal Clinically Important Difference

### G. Feedback and Other Interventions

We found three studies (fair quality; Table 5B) that assessed the effects of different interventions on UL motor impairment and activity levels in individuals post-TBI. Sietsema and colleagues^54^ assessed the effects of playing a game within an occupational context compared to rote exercises on UL movement patterns. Twenty individuals (17 males, 3 females) with mild to moderate TBI participated in the study. Participants practiced 10 trials in both conditions. The total forward reaching distance from the hip to the wrist was measured using motion analysis. Game playing resulted in greater reaching distance (13cm more, ES=0.63) than rote arm reaching exercises.

Croce and colleagues^55^ evaluated the effectiveness of provision of knowledge of results (KR) feedback at different schedules in subjects with severe TBI (n=51; 42 males, 9 females). All participants practiced 60 trials (5 trials/block, 12 blocks) of an anticipation task. Participants received KR feedback on timing errors after each trial at different schedules – no KR (n=12), 100% KR (n=14), summary KR (n=13) and average KR (n=12). They were then tested for immediate (after 10 minutes) and delayed (after one hour) retention. All the three KR groups were more accurate in the last block compared to the first block of trials (ES=0.96). At early retention testing, this effect was decreased in the 100% KR group. However, the summary KR (ES=1.21) and average KR (ES=1.02) groups continued to improve accuracy. At the late retention testing, the effects were further reduced in the 100% KR group (ES=0.37) and average KR group (ES=0.77) but was retained only in the summary KR group at the same level (ES=1.21).

Sterr and Freivogel^56^ examined the effects of shaping principles on UL activity performance in 13 individuals (9 males, 4 females) with ABIs (TBI: n=11, stroke: n= 2). All participants were evaluated using the MAL, WMFT and Frenchay Arm Test. Compared to provision of Occupational Therapy, task-practice using shaping principles resulted in greater motor improvement on all outcomes (MAL; AoU: ES=2.23, QoM: ES=1.98), WMFT; ES=1.76 and the Frenchay Arm Test; ES=0.72).

## Discussion

We examined the effectiveness of different interventions to augment UL motor improvement in individuals post-TBI. Majority of the studies reported moderate to large effect sizes for intervention effectiveness. In terms of quality assessment, one study was excellent, five good and the rest were fair.

### Outcomes used to assess motor improvement

A variety of outcomes were used to assess motor improvements at different levels of the International Classification of Functioning (ICF).^57^ At the motor impairment level, the FMA was the used most commonly.^41, 42, 48, 51, 53^ Goniometry^34, 35, 49^ and torque controlled passive extension^37^ helped assess changes in wrist and elbow ranges of motion. In addition, kinematic motor performance outcomes including speed, reaching path straightness and accuracy helped quantify motor impairment.^44–46^ These kinematic measures were obtained using motion capture equipment, robotic manipulandum or using instrumented tablets. All of the above-mentioned measures have well established psychometric properties.^58, 59^

Muscle tone was most commonly quantified using the Ashworth’s scale or the MAS.^34, 35, 38^ Other measures used included the Modified Tardieu Scale^37^ or neurophysiological (H-Reflex) measures.^39^ The MAS has been recommended as a measure of choice in published guidelines.^58^ However, both the MAS and Modified Tardieu Scale have poor inter-rater reliability in individuals post-TBI.^60^ Use of the MAS alone does not distinguish between the tonic and phasic components of spasticity.^61^ Changes noted in H-reflex based parameters do not automatically translate to better functional performance after rehabilitation.^62^ The utility of other neurophysiological measures (e.g. based on spatial threshold control of muscle activation^61^ alone or in conjunction with existing clinical measures to assess muscle tone remains to be estimated.

Similar to motor impairment, a variety of assessments were used to measure activity limitations. The WMFT was used most commonly ^40–42, 56^ across the different studies. Dexterity was measured by using the BBT,^46, 48, 51^ Purdue Pegboard Test,^43^ TEMPA^37, 52^ and Jebsen Taylor Hand Function test^48^ in different studies. Limitations in ADL performance were also quantified using the FIM,^36, 53^ the CHART,^36^ Frenchay Arm Test^56^ and the ARAT.^40^ In addition, studies also used the DASH^47^ MAL amount and quality scores,^40–42, 56^ and the Stroke Impact Scale^51^ which report participant self-perceived levels of UL use. All the measures have excellent psychometric properties^59^ and the FIM and ARAT are part of the published guidelines.^58^ Inclusion of the DASH, MAL and Stroke Impact Scale across studies is encouraging, given the suggestion to use patient reported measures as outcomes in intervention studies.^63^

### Follow-up assessments

It has been suggested that motor improvement after TBI is attributable in part to motor learning.^12^ Retention of improvements in performance noted at the end of the intervention denote motor learning. However, only 10^36–39, 41, 44, 51, 52, 55, 56^ studies included any form of retention testing. Amongst these studies, the timing of testing varied widely. Retention was tested at the following periods post-intervention: 10 minutes,^55^ 20 minutes,^39^ 30 minutes,^44^ one hour,^38^ three hours,^50^ 24 hours,^37, 50, 55^ four weeks,^37, 41, 56^ six weeks,^36^ six months,^51^ one year^52^ and two years post-intervention.^41^

Not all studies found that changes were retained. While hypertonia was reduced in the short-term (≤24 hrs) using casting^37^ and acupuncture,^39^ long-term retention (>24 hours) was absent with oral tizanidine.^36^ Only short-term retention was assessed with VR^44^ and feedback provision.^55^ Use of shaping principles with^41^ and without^56^ constraint as well as Arm Motor Ability Training^52^ resulted in long-term retention. Both short^50^ and long-term^51^ retention were seen with the use of tDCS. It remains to be seen if use of VR technology and use of different interventions including acupuncture and Botulinum toxin A result in long-term retention in individuals post-TBI.

### Presence of cognitive and mood impairments

Dysfunction in different cognitive domains influences generalized motor improvement in individuals post-TBI.^6^ Only two^42, 50^ of the selected studies, examined the association between UL motor improvement and cognitive impairment. Few other studies provided information on baseline levels of cognitive functioning,^40, 48, 54, 55^ but did not examine the effects of baseline cognitive dysfunction with motor improvement. Only one study assessed the levels of baseline depressive symptomatology,^48^ which can predict motor improvement and satisfaction with life after discharge from rehabilitation in this population.^64^ The presence of cognitive impairments^65^ and depressive symptoms^66^ influence motor improvement after a stroke. Future studies will need to focus on the relationship between cognitive dysfunction, mood disorders and motor recovery in individuals post-TBI to better understand their association with motor improvement.

### Level of injury severity

Out of the 23 included studies, only few studies reported initial injury severity levels. The Glasgow Coma Scale,^34, 35, 37–39, 53, 55^ duration of post-traumatic amnesia^45^ or Rancho Los Amigos original scale^54^ helped quantify initial injury severity levels. This information is an important prognostic indicator for changes in overall motor improvement and levels of activity performance assessed using the Barthel Index^67^ as well as a composite score of activities of daily living and social participation (assessed using the Glasgow Outcome Scale Extended measure).^68^

The other studies did not specify the injury severity levels, but some provided FMA scores.^41, 42, 44, 45, 47, 51^ FMA scores ≥50/66 and ≤49/66 represent as mild and moderate-to-severe levels of post-stroke UL motor impairment.^69^ The FMA scores in the acute post-stroke stage can predict subsequent UL recovery potential.^70^ Whether UL FMA scores can be used to make similar predictions in individuals post-TBI remains to be estimated.

### Sex and gender considerations

As stated previously, a greater proportion of males (1.2-4.4) sustain TBIs compared to females.^3,71^The greater proportion of males amongst the included participants across the different studies are indicative of the above findings. Only two studies had an almost equal distribution of sexes,^46^ or included more females.^44^ Despite consistent calls for considerations of sex and gender on functional outcomes,^72, 73^ only one study^47^ assessed the effects of sex on outcomes. Future studies must strive to include more female participants and consider the effects of sex and gender on functional outcomes.

### Effect of chronicity

Studies on interventions including acupuncture, CIMT, VR-based games, NMES, stem-cells, game-playing, feedback, and forced-use therapy exclusively included participants with chronic injuries. While studies using Botulinum toxin A included acute, sub-acute and chronic participants, separate analyses were conducted by chronicity.^34^ Use of serial-casting^37^ included only participants in the sub-acute stage. Other studies including participants across all stages did not conduct-separate analyses based upon chronicity.^38, 50, 52^ Future studies must strive to include participants across all stages or conduct sub-analyses based on time since injury.

## Limitations

Heterogeneity amongst the interventions used prevented performance of an overarching statistical synthesis like a meta-analysis. Amongst the 23 studies included in this review, only 9 studies were designed as RCTs. Although the wide variability in presenting symptoms and underlying injury severity present serious challenges in designing RCTs involving individuals post-TBI,^74^ encouraging efforts are underway.^75^ Only three studies included in this review had sample size calculations^37, 45, 52^ and one study^48^ provided an estimate for numbers of participants needed for future trials. Nine of the 23 studies included participants with stroke and TBI. Thus, generalization of the findings is limited to a certain extent, except for two studies,^47, 51^ which conducted separate analyses for individuals post-TBI. Future studies should include only individuals post-TBI or conduct separate sub-analyses for this population. Limits placed on age (adults), language (English only) and non-inclusion of terms like shoulder, elbow, wrist, hand, etc. may have led to exclusion of some studies.

## Conclusion

Preliminary evidence suggests that different rehabilitation interventions may facilitate UL motor improvement in individuals post-TBI. This systematic review has identified several new questions in individuals post-TBI including whether provision of: i) Botulinum toxin A followed by intensive rehabilitation results in better long-term reduction of muscle tone; ii) CIMT results in better motor improvement compared to traditional therapy; iii) a combination of interventions such as VR-based gaming and tDCS is more beneficial than provision of one single intervention; and iv) provision of knowledge of performance feedback is useful and results in similar or better improvements than KR feedback. We hope that these questions will help guide and foster further research to evaluate the efficacy of the most suitable interventions to reduce impairment and improve activity performance post-TBI.

## Supporting information

Downs and Black scoring

## Data Availability

The search strategy is outlined in the paper.

## Acknowledgements

The authors would like to acknowledge Dr. Kate Aultman for her support and encouragement in this project.

## Abbreviations

TBI: Traumatic Brain Injury
UL: Upper Limb
LL: Lower Limb
PICO: Patient, Intervention, Comparison, Outcome
MKF: Melinda K Fountain (2^nd^ author)
AFH: Ashley F Hood (3^rd^ author)
SKS: Sandeep K Subramanian (1^st^ author)
ES: Effect Size
MAS: Modified Ashworth’s Scale
ABI: Acquired Brain Injury
CIMT: Constraint Induced Movement Therapy
MAL: Motor Activity Log
FMA: Fugl-Meyer Assessment
WMFT: Wolf Motor Function Test
BBT: Box and Blocks Test
BBT: Box and Block Test
DASH: Disabilities of the Arm, Shoulder and Hand
BBS: Berg Balance Scale
NMES: NeuroMuscular Electrical Stimulation
FIM: functional Independence Measure
ICF: International Classification of Functioning, Disability and Health
CHART: Craig Handicap Assessment and Reporting Technique
ARAT: Action Research Arm Test

